# Plasma Proteomics and Diabetes Duration Predict Aneurysm Incidence and Rupture Using Machine Learning

**DOI:** 10.1101/2025.02.25.25322902

**Authors:** Bichen Ren, Tingfeng Xu, Lu Yu, Xingyu Chen, Xiong Yang, Yuan Fang, Tianyue Pan, Hongqiang Zhao, Jichen Wen, Penghui Wang, Liang Chen, Gang Fang, Weiguo Fu, Jiucun Wang, Minxian Wang, Zhihui Dong

## Abstract

**Background:** Early identification of individuals at high risk for aneurysms, particularly ruptured aneurysms, is critical for timely intervention. However, existing imaging-free prediction models have significant limitations. This study aims to develop a robust model for predicting aneurysm incidence and rupture by leveraging multi-omics data, including circulating proteomics, and identifying specific biomarkers.

**Methods:** UK Biobank participants (n = 502,389; mean age: 58.0 years; 54.4% female) without a history of aneurysms were divided into a training set, which included a derivation set (n = 473,630) and a validation set (n = 8,628), as well as a testing set (n = 20,131) for model evaluation. Cox proportional hazards (CPH) models were used to estimate the risk of aneurysm events, including total, unruptured, and ruptured aneurysms across three types: aortic aneurysm (AA), abdominal aortic aneurysm (AAA), and intracranial aneurysm (IA). We developed base models that incorporated plasma proteomics (Proteins), metabolomics (Metabolites), polygenic risk scores (PRS), and clinical risk factors (RF) to predict nine aneurysm-related outcomes using 10-fold cross-validation with LASSO regression. Additionally, we investigated the relationship between diabetes duration and aneurysm events and developed a classification model, the Diabetes Duration Score (DDscore), to enhance model performance.

**Results:** During the 14.8-year follow-up, there were 4,292 AA events, 2,730 AAA events, and 3,644 IA events. The Proteins Model demonstrated superior or comparable discriminative performance for most AA and AAA endpoints, with C-indexes exceeding 0.9 for rupture events. However, no predictive advantage was observed for IA endpoints. For different time windows, the Proteins Model achieved the highest AUC for most endpoints within 5 years. Time-dependent analysis revealed an opposing relationship between diabetes duration and aneurysm risk: shorter diabetes duration was associated with higher risk, while longer duration reduced risk. Adding DDscore significantly improved predictions for AA and AAA, particularly for ruptured AAA (C-index [95% CI]: Proteins + DDscore 0.93 [0.88-0.99] and Proteins + RF + PRS + Metabolites + DDscore 0.94 [0.91-1.00]). For clinical utility, the Proteins or Proteins + DDscore models provided greater net benefit at low decision thresholds (0%-2% for ruptured AA and 0%-1% for ruptured AAA). Additionally, 30 rupture-specific plasma proteins with high weight were identified for all types of aneurysms.

**Conclusions:** Plasma proteomics and diabetes duration demonstrated exceptional predictive capabilities for aneurysm events, particularly rupture. The machine-learning model developed in this study achieved accurate predictions even up to 10 years before diagnosis, with potential implications for high-risk screening and early intervention.

## Introduction

Aneurysms are pathological localized dilatations of arteries, classified by location into various types such as intracranial, aortic, abdominal, and peripheral aneurysms, with a prevalence of 0.92% to 3.2%^1–3^. The most severe complication is rupture, which can cause sudden hemorrhage and high mortality. Ruptured aortic aneurysms (AA) and abdominal aortic aneurysms (AAA) have a pre-hospital fatality rate of 60-80%^4,5^, with AAA rupture survival rates below 50% even with immediate surgery^6^. Rupture of intracranial aneurysms (IA) results in poor prognosis despite some survival^7^. Early identification of high-risk individuals is essential to allow timely intervention, especially for rupture-prone aneurysms.

Aneurysms often remain asymptomatic, growing slowly until rupture, complicating early detection. Global screening aims to identify aneurysms or arterial dilation at early stages^4,8–11^. Several risk stratification models based on clinical predictors exist^12,13^, yet uncertainties remain. For instance, diabetes’s role—protective or risk factor—in aneurysm development is still debated, potentially impacting prediction accuracy^14–16^. Genetic predisposition is significant, with aneurysm heritability estimated at 70% for AAA and 41% for IA^17,18^. Genome-wide studies have identified 24 loci for AAA and 17 for IA^17–20^. Traditional predictions largely rely on clinical factors and polygenic risk scores (PRS), but predicting rupture remains challenging due to the genetic similarity between ruptured and unruptured IAs^18^. Effective rupture prediction primarily depends on short-term imaging follow-ups for aneurysm growth or morphometric machine learning models using Computed Tomography Angiography (CTA)/Magnetic Resonance Imaging (MRI) data. However, most studies use surrogate markers (e.g., accelerated growth or instability) rather than direct rupture endpoints^21,22^, complicating prediction for undetected or asymptomatic aneurysms lacking imaging data.

Recent advances in blood biomarkers offer a promising alternative, especially for aneurysm rupture prediction within the general population. In this study, we adopted a data-driven, multi-modality approach using a large prospective cohort with extended follow-up to identify molecular features associated with aneurysm incidence or rupture risk and evaluate their predictive performance. Leveraging comprehensive UK Biobank data on over 500,000 participants, including plasma proteomics, NMR metabolomics, genomics, and clinical predictors, to address the following objectives: (1) to systematically evaluate the predictive power of various modalities features for predicting aneurysm events based on machine-learning models; (2) to examine the relationship between diabetes and aneurysms to enhance model robustness; (3) to assess model accuracy, clinical utility, and optimal prediction windows; and (4) to pinpoint specific blood biomarkers associated with aneurysm events and quantify their contributions.

## Methods

### Study population and endpoint definition

We use data from the UK Biobank (UKB) cohort, and all participants were enrolled from 2006 to 2010 in 22 recruitment centers across the United Kingdom and had a median follow-up of 14.8 years until January 2024. We investigated a set of nine endpoints for aneurysms, each defined by the earliest occurrence in primary care after enrollment, including hospital episode statistics or death records. Endpoints defined by ICD codes (**Table S1**) and group information (**Figure S1**), and individuals with prevalent cases (first occurrence before or up to the baseline assessment visit) or incident cases recorded within the first month of follow-up were excluded for each endpoint. Aneurysm diseases are categorized into three types: aortic aneurysms (AA), abdominal aortic aneurysms (AAA), and intracranial aneurysms (IA). AA includes AAA, thoracic aneurysm, and thoracoabdominal aortic aneurysm, regardless of whether they cross the diaphragm, due to the limited number of cases affected by ICD in the UKB showcase. The endpoint events for each type of aneurysm are defined as all, non-ruptured, and ruptured. If there is a record of aneurysm rupture, the participant is recorded in the rupture subgroup and excluded from the unruptured subgroup.

### Development and interpretability of single- and multi-modality models

The UK Biobank Plasma Proteomics Project (UKB-PPP) utilized the Olink Explore 1536 and Explore Expansion platforms to conduct plasma proteomic profiling, targeting 2,923 unique proteins across 2,941 assays^25^. Following the filtering process, data normalization was applied to a total of 2,911 unique proteins. The Proteins Model was developed using a 10-fold least absolute shrinkage and selection operator (LASSO) regression approach, targeting 9 aneurysm events, with a derivation set of N = 24,023 individuals, under Python using cuML (v24.6.1)^26^.

For Metabolites Model development, only the 169 original measures were included after filtering out any samples with missing measurements and applying data normalization. We implement the Metabolites Model by employing a 10-fold LASSO regression model for 9 endpoints utilizing the cuML package (v24.6.1)^26^, with a derivation set of N = 232,221 individuals.

Polygenic risk scores (PRS) for thoracic aortic aneurysm^27^ and IA^18^ were derived using LDpred2 algorithm^28,29^, while PRS for AAA^30^ was derived using a previously published PRS based on Bayesian regression with continuous shrinkage priors (PRScs)^31^. The genome-wide association study (GWAS) summary statistics used to derive the PRS were sourced from European populations and were non-overlapping with the UK Biobank to avoid potential bias. Ultimately, for the 9 endpoints related to AA, AAA, and IA, each PRS was used to construct a predictive PRS Model. (**Table S2**).

The study highlights 12 key clinical risk factors (RF) for aneurysms, including age, sex, BMI, low-density lipoprotein (LDL) cholesterol, high-density lipoprotein (HDL) cholesterol, triglycerides, total cholesterol, current smoking, systolic blood pressure (SBP), diastolic blood pressure (DBP), atherosclerotic disease (coronary artery disease, peripheral artery disease, and ischemic stroke), family history of CVD.

The Multi-modality Models were developed using a linear regression approach to ensemble scores from a validation set with comprehensive and normalized omics data using the package scikit-learn (v1.5.1)^32^. The weights of each single-modality profile in the Multi-modality Model were derived from the validation set, and overall performance was assessed using the testing set.

The LASSO regression model’s interpretability stems from its feature selection capability. To evaluate the model’s interpretability, we analyzed the non-zero coefficients obtained after fitting the LASSO model to data from prediction models for 9 aneurysm events. We applied Max-Normalization to these coefficients and visualized the identification of aneurysm-specific proteomic and metabolite profiles, as detailed in **Table S3 and S4**.

### Development of the Diabetes Duration Score classification model

To explore the relationship between diabetes and aneurysm events, we assessed the correlation between nine endpoints and diabetes-related features, including prevalent diabetes, incident diabetes, antidiabetic medication, HbA1c level, and glucose, using Cox regression, adjusting for age, sex, BMI, smoking status, assessment center, lipid-lowering medications, antihypertensive drugs, and genetic principal components (**Table S5**). P values reported in forest plots are using a two-sided Wald test and values less than 0.001 were considered significant.

Furthermore, diabetes duration was defined as the time of diabetes diagnosis to aneurysm events, which was used as an important time variable for relationship studies. The correlation was evaluated using Cox regression as the primary analysis, and the 11 adjustment factors have been increased to include antidiabetic medication, HbA1c, and glucose levels. Based on the time-dependent correlations between diabetes and aneurysm events, we segmented diabetes duration using specific time cut-off points and classified it into four groups: non-diabetes, short-term diabetes, medium-term diabetes, and long-term diabetes, corresponding to each aneurysm event (**Table S7-S10**). We developed a time-to-event classification model named Diabetes Duration Score (DDscore) for each aneurysm endpoint. DDscore utilizes normalized clinical predictors (**Table S11**) processed through the Extreme Gradient Boosting (XGBoost) algorithm^33^, with hyperparameter tuning performed using Ray Tune^34^ on the derivation set (**Table S12**). Due to practical application, the DDscore in the validation and testing set was imputed with results obtained from the based model. To ensemble with the single- and multi-modality models, we developed two further models: one that integrated Proteins and DDscore; and another that incorporated Proteins, RF, PRS, Metabolites, and DDscore. Linear regression was used on the validation set, and predictive performance was assessed on the testing set.

### Statistical analysis

We applied Cox proportional hazards (CPH) model^35^to predict risks for nine aneurysm endpoints. Model development was independently conducted for each derivation set. For every cross-validation split, models were trained on the respective training sets, and model checkpoints were selected using the validation sets. CPH models were fitted using the CoxPHFitter from the Python package lifelines (v0.27.8)^36^,We assessed these models using Harrell’s C-index over 100 bootstrap samples, where the index ranges from 0.5 for noninformative models to 1 for perfectly discriminating models. Final evaluations aggregated predictions on the testing set.

Calibration plots were utilized to visually present the alignment between predicted risks and observed event rates. Calibration was evaluated using the Brier score, which calculates the mean squared error between actual outcomes and predicted probabilities^37^using the scikit-learn package (v1.5.1). Net benefit curves were also drawn using the dcurves package (1.0.6.5) under Python to illustrate the different predictive contributions of each model.

We reported several predictive metrics, including the area under the Receiver Operating Characteristic (ROC) curve (AUC), the area under the precision-recall curve (APR), as well as metrics like accuracy, sensitivity, and specificity. based on the cut-off corresponding to the largest Youden index. The predictive performance of the models was evaluated across three-time intervals: within 5 years, 10 years, and beyond 10 years. Events occurring beyond 10 years were treated as non-incidents for the 5- and 10-year analyses, and participants with events within 10 years were excluded from the beyond-10-year evaluation.

Detection rates (DR) and likelihood ratios (LR) were computed in the testing set at a false positive rate (FPR) of 10%. FPR was calculated using the formula: FPR = false positives (FP) / (true negatives (TN) + FP), while DR was calculated as DR = true positives (TP) / (false negatives (FN) + TP). LRs were then determined as LR = DR/FPR.

We calculated category-free net reclassification improvements (NRI) by incorporating the DDscore into both single- and multi-modality models, using the largest Youden index as a threshold to ensure conservative estimates of risk difference. Additionally, we computed integrated discrimination improvements by adding the DDscore to the single- and multi-modality models using R.

All statistical analyses and modeling were performed using Python (3.10.14) and R software (4.3.3). P < 0.001 was considered significant.

## Results

### Study population and dataset information

This study adopted 502,389 participants from 22 centers with clinical and multi-modality data currently available in the UK Biobank (UKB), and a prospective cohort of median enrollment age of 58.0 years (interquartile range [IQR] 50–63), of whom 54.4% were female, and mainly consisted of white ethnicity (78.7%) (**Table 1**). Body mass index was 26.7 (IQR 24.1–29.9), systolic blood pressure was 139.0 mmHg (IQR 126.0–155.0), and 52,944 (10.5%) people were current smokers. Excluding participants with a history of aneurysms within 30 days of enrollment, we observed 4,292 AA, 2,730 AAA, and 3,644 IA incident cases. Among these, aneurysm rupture occurred in 318 (7.4%), 259 (9.5%), and 3,132 (86.0%) participants, respectively, over a median follow-up of 14.8 years (IQR 14.0–15.5) until January 2024 (**Table 1, Figure S1**). We extracted clinical predictors and aneurysm events for all individuals with multi-modality profiling during cohort recruitment. The three types of aneurysms each include three endpoints: aneurysm, unruptured aneurysm, and ruptured aneurysm, totaling nine endpoints. These profiling measurements included 487,185 genomic, 53,020 proteomic, and 260,976 NMR metabolite datasets (**Figure 1a**). A total of 20,131 individuals were included in the testing set, which was a randomly selected 70% of the population with complete clinical and multi-modality data. The remaining 30% of individuals used to validate the integration model were included in the training set along with others possessing complete basic information, totaling 482,258 participants (**Figure 1b**, **Figure S2**). After model selection on the validation datasets and obtaining the selected models’ final predictions on the testing set for downstream analysis (**Figure 1c**).

**Table 1.** Baseline Characteristics of UK Biobank participants in the study.

|  | **All participants**<br>**(N = 502,389)** | **Training set**<br>**(N = 482,258)** | **Testing set**<br>**(N = 20,131)** |
| --- | --- | --- | --- |
| Age at recruitment | 58.0 [50.0,63.0] | 58.0 [50.0,63.0] | 58.0 [50.0,64.0] |
| Sex (male) | 228982 (45.6) | 219670 (45.6) | 9312 (46.3) |
| Ethnicity (white) | 395387 (78.7) | 379095 (78.6) | 16292 (80.9) |
| Body mass index, kg/m<sup>2</sup> | 26.7 [24.1,29.9] | 26.7 [24.1,29.9] | 26.8 [24.2,30.0] |
| Systolic blood pressure, mmHg | 139.0 [126.0,155.0] | 139.0 [126.0,155.0] | 140.0 [126.0,155.0] |
| Diastolic blood pressure, mmHg | 83.0 [75.0,91.0] | 83.0 [75.0,91.0] | 83.0 [75.0,91.0] |
| LDL cholesterol, mmol/L | 3.7 [3.2,4.3] | 3.7 [3.2,4.3] | 3.7 [3.2,4.3] |
| HDL cholesterol, mmol/L | 1.4 [1.2,1.7] | 1.4 [1.2,1.7] | 1.4 [1.2,1.7] |
| Triglycerides, mmol/L | 1.5 [1.1,2.2] | 1.5 [1.1,2.2] | 1.5 [1.1,2.2] |
| Total Cholesterol, mmol/L | 5.8 [5.1,6.6] | 5.8 [5.1,6.6] | 5.8 [5.1,6.6] |
| Current Smoker | 52944 (10.5) | 50900 (10.6) | 2044 (10.2) |
| History of coronary artery disease | 21086 (4.2) | 20052 (4.2) | 1034 (5.1) |
| History of peripheral artery disease | 3506 (0.7) | 3301 (0.7) | 205 (1.0) |
| History of ischemic stroke | 3318 (0.7) | 3156 (0.7) | 162 (0.8) |
| History of diabetes | 13559 (2.7) | 12867 (2.7) | 692 (3.4) |
| History of chronic kidney disease | 2281 (0.5) | 2129 (0.4) | 152 (0.8) |
| History of atrial fibrillation | 28609 (5.7) | 27242 (5.6) | 1367 (6.8) |
| History of heart failure | 15625 (3.1) | 14838 (3.1) | 787 (3.9) |
| History of hypertension | 136157 (27.1) | 130332 (27.0) | 5825 (28.9) |
| Family history of CVD | 375899 (74.8) | 360713 (74.8) | 15186 (75.4) |
| Cholesterol treatment | 87077 (17.3) | 83201 (17.3) | 3876 (19.3) |
| Hypertension treatment | 2111 (0.4) | 2016 (0.4) | 95 (0.5) |
| Follow-up time, years | 14.8 [14.0,15.5] | 14.8 [14.0,15.5] | 14.8 [14.1,15.6] |

Continuous data are presented as median [interquartile range] and categorical variables as number (percentage). LDL, low-density lipoprotein; HDL, high-density lipoprotein; CVD, cardiovascular disease.

**Figure 1.**
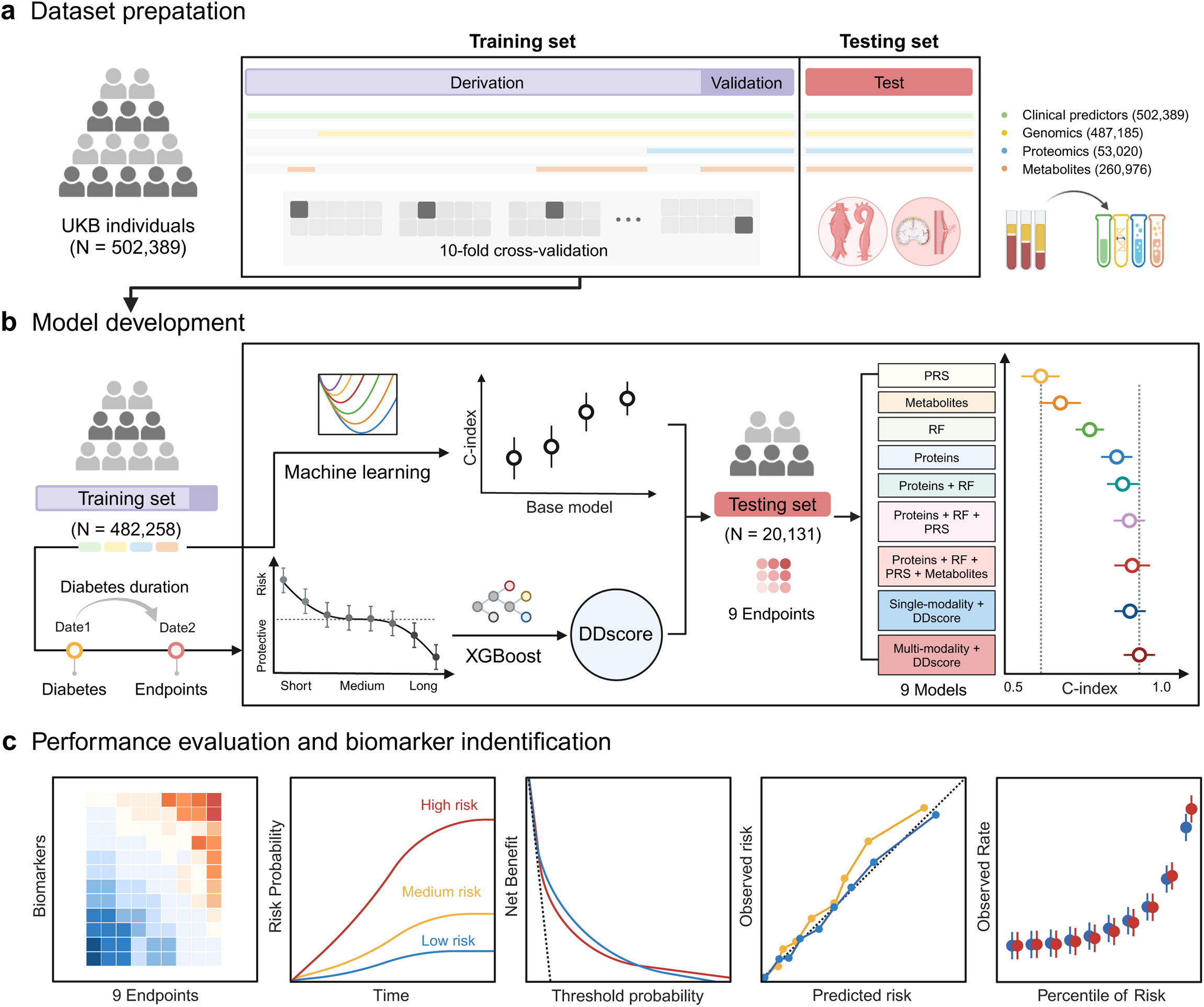
Study overview. **a,** Among UKB participants (N = 502,389) with a median follow-up time of 14.8 years, 28,759 individuals have complete multi-modality data, including clinical predictors, genomics, proteomics, and metabolomics. Randomly divide these individuals into validation (N = 8,628) and testing sets (N = 20,131) at a 3:7 ratio. The remaining 473,630 individuals, who have 1-3 types of information data, will serve as the Derivation set (N = 473,630), constituting the training set with the validation set. **b,** In the training set, the base model was developed to predict 9 aneurysm endpoints, respectively using 12 clinical risk factors, aneurysm-associated PRSs, 2,911 plasma proteins, and 169 metabolomic markers with a 10-fold cross-validation LASSO regression. Based on the time-dependent relationship between diabetes duration and aneurysm events found, a classification model called DDscore was developed on the training set using XGBoost. Compare the discrimination of the integrated model obtained through DDscore with that of the baseline model. **c,** The model performance evaluations in stratifying at-risk populations aimed to inform their calibration, discrimination, and clinical usefulness, and to identify the biomarkers with high weights and strong specificity within the population to explain machine learning models. UKB, UK Biobank; PRS, polygenic risk score; RF, clinical risk factor; DDscore, diabetes duration score; LASSO, least absolute shrinkage and selection operator; XGBoost, extreme gradient boosting. Created with BioRender.com.

### Base models for single- and multi-modality data

Many clinical risk factors (RF) are accessible in primary care and are commonly used to stratify aneurysm risk^9,12^. However, achieving good predictive performance often requires complex risk-scoring systems and methods. We constructed baseline models using RF, PRS, Metabolites, and Proteins to predict nine aneurysm endpoints. These models were trained with a 10-fold cross-validation LASSO regression, incorporating 12 key clinical risk factors (details in **Methods**), aneurysm-related PRS (**Table S2**), 2,911 plasma proteomic profiles (**Table S22**), and 169 metabolomic markers (**Table S3**), all available in the UKB. We modeled aneurysm event risk for each endpoint using Cox proportional hazards (CPH) models for four baseline models. Multi-modality Models were developed using a linear regression approach with increasing complexity to ensemble scores from the validation set. For the testing set, the discriminative performances of all models for all events were quantified by Harrell’s C-index. In most AA and AAA endpoints, the Proteins Model had significantly greater or comparable discriminative performance than RF, PRS, and Metabolites Models. Notably, in AA and AAA rupture events, the C-indexes of the Proteins Model exceeded 0.9, indicating that Proteins generally provide more competitive predictive information than other models (**Figure 2a**). However, no predictive advantage of Proteins was observed in IA endpoints, where the RF Model demonstrated better discriminative performance. Multi-modality models were integrated to achieve the best predictive model, with the Proteins + RF + PRS + Metabolites Model showing the greatest C-index. Discriminative performances were significantly improved, especially in unruptured AA and AAA endpoints. Compared to the PRS, the Proteins Model significantly improved classification ability for all aneurysm events (**Figure 2b, Table S6**). Notably, the likelihood ratio (LR) improvement was noteworthy for AA and AAA, especially for ruptured AA (ΔLR = 5.95) and ruptured AAA (ΔLR = 4.32). However, the Proteins + RF + PRS + Metabolites slightly increased LR than the Proteins Model.

**Figure 2.**
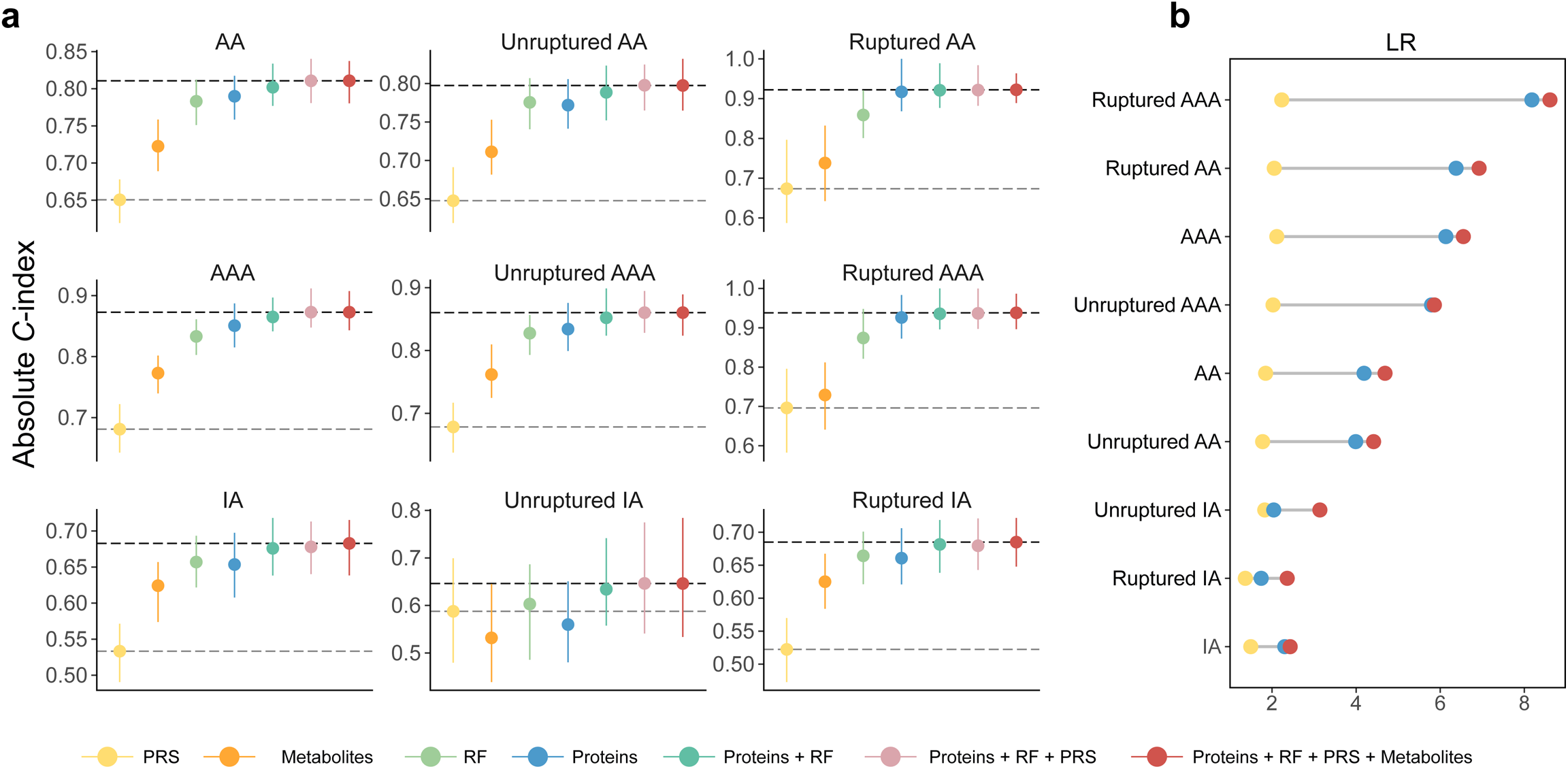
Predictive value and enhanced performance of single- and multi-modality models for aneurysm events. **a,** Comparison of discriminative performance of CPH model performance trained on PRS (yellow), Metabolites (orange), RF (light green), Proteins (blue), Proteins + RF (dark green), Proteins + RF + PRS (pink), and Proteins + RF + PRS + Metabolites (Four-modality, red). Differences in performance between the PRS baseline (gray dashed line) and the Four-modality model (black dashed line) are shown. Data are presented as medians (error bar centers) with 95% CIs (error bar lines) from 100 bootstrap iterations (Table S16). **b,** Comparison of LR (ΔLR, gray line) at 10% FPR for PRS (yellow), Proteins (blue), and the Four-modality model (red) (Table S6). LR, likelihood ratio; FPR, false positive rate.

Participants with a higher percentile of model risk score at baseline exhibited elevated observed event rates across almost all aneurysm endpoints (**Figure S3a**). The Kaplan-Meier survival curves showed distinctive paths between the tertiles stratified by the Proteins Model (**Figure S3b, Table S13**). Compared with the bottom tertile, individuals with top-tertile protein levels have over six times the risk of AA rupture (hazard ratio (HR) [95% confidence interval (CI)]: 6.34 [4.29–9.38]) and AAA rupture (HR: 7.45 [4.83–11.50]). The HR for the Proteins + RF + PRS + Metabolites Model is similarly elevated. Furthermore, the top decile of Proteins exhibited a higher prevalence of ruptured AA than the Proteins + RF+ PRS + Metabolites Model (**Figure S4a**). We investigated the predictive abilities across different time windows; for most endpoints, the Proteins Model achieved the highest area under the ROC curve (AUC) when forecasting outcomes happening within 5 years, suggesting a pivotal role of plasma proteomics in detecting near-term risks (**Table S14**).

### Time-dependent correlation of diabetes and aneurysm events

So far, guidelines and screening recommendations for aneurysms have not recognized diabetes as either a risk or protective factor. Nevertheless, various studies have explored how diabetes affects the development and progression of aneurysms, revealing contradictory effects that are risky, protective, or neutral^14–16^. In the UKB cohort, 46,791 participants were diagnosed with diabetes during the follow-up period (**Table S15**). Compared to participants without diabetes, they showed a higher proportion in unruptured AA (1.7%) and AAA (1.2%), as well as in ruptured IA (1.1%). To explore the relationship between diabetes and aneurysm events, we assessed the correlation between nine endpoints and diabetes-related features, including prevalent diabetes, incident diabetes, antidiabetic medication, glycated hemoglobin (HbA1c) level, and glucose. Diabetes incidence after enrollment significantly increased the risk of most aneurysm endpoints, except AA and AAA rupture. Still, diabetes at enrollment was not the protective factor due to the wide confidence intervals (CIs) (**Table S5**). Hyperglycemia and the use of antidiabetic medications as potential protective factors for aneurysm events did not show consistent results. None of the diabetes-related variables could be identified as protective factors for IA. The relationship between HbA1c level and glucose with endpoints was contradictory or not statistically significant for AA and AAA, making it difficult to determine the impact of hyperglycemia.

We hypothesize that these inconsistent correlations are attributed to the timing of diabetes’s influence on aneurysms. Therefore, diabetes duration was defined as the time of diabetes diagnosis to aneurysm events. The time-dependent correlations between diabetes and aneurysm events were found using Cox regression after adjustment. We observed a similar and stable pattern of association between diabetes duration and all aneurysm endpoints: a shorter duration of diabetes was associated with fewer events, whereas a longer duration of diabetes led to increased events; in contrast, the events during the intermediate duration showed a relatively fluctuating pattern. (**Figure 3a, Table S7**). In the sensitivity tests, we matched participants in diabetes and non-diabetes using propensity score matching (PSM) at three different ratios based on all covariates from the primary analysis, and the pattern of time-dependent correlation remained unchanged **(Table S8-S10)**.

**Figure 3.**
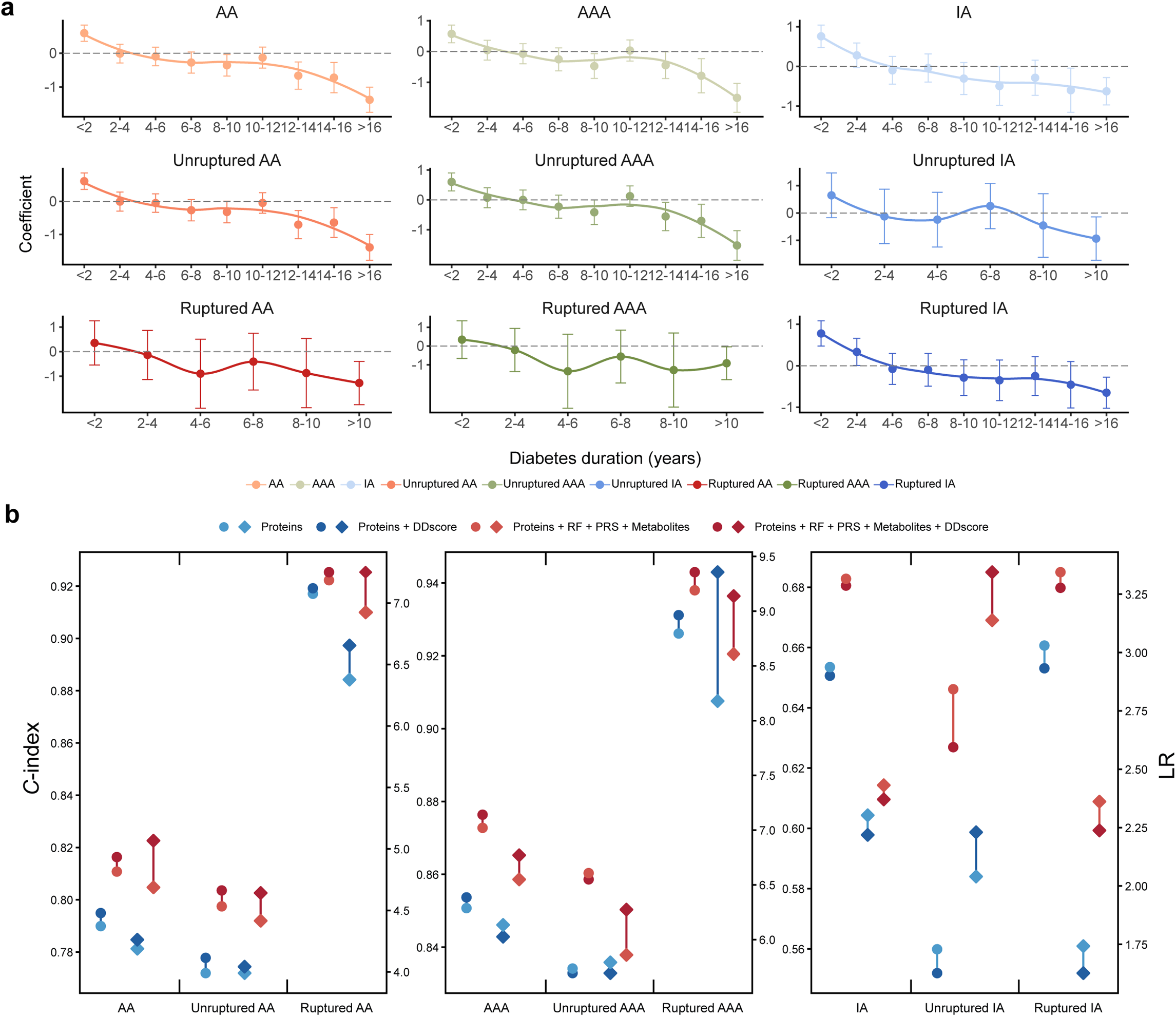
Enhancement in the predictive model through time-dependent correlation between diabetes and aneurysm events. **a,** The correlations between diabetes duration and nine aneurysm events were evaluated using Cox regression (Table S7). Diabetes duration, the time of diabetes diagnosis to aneurysm events. **b,** Comparison of the discriminative performance of CPH models trained on Proteins (blue), Proteins + DDscore (dark blue), Proteins + RF + PRS + Metabolites (Four-modality, red), and Proteins + RF + PRS + Metabolites + DDscore (dark red); Circles represent the C-index values on the left, and diamonds represent the LR values on the right; The addition of DDscore improved the C-index over the benchmark base model (Table S16) and enhanced LRs at a 10% FPR (Table S6); DDscore, a time-to-event classification model named Diabetes Duration score; LR, likelihood ratio; FPR, false positive rate.

### Discriminative improvements of the model integrating the Diabetes Duration score

Based on the time-dependent correlations of diabetes and aneurysm events found, we categorized diabetes duration into four groups: non-diabetes, short-term, medium-term, and long-term diabetes. We developed a time-to-event classification model named Diabetes Duration score (DDscore) for each aneurysm endpoint through the Extreme Gradient Boosting (XGBoost) algorithm using normalized clinical predictors (**Table S11-S12**). To integrate with the single- and multi-modality models, we developed two further models, the Proteins + DDscore Model, and the Proteins + RF + PRS + Metabolites + DDscore Model, to assess predictive performance on the testing set. The feature standardized weights after model fitting were consistent with the previous results, including the Proteins with the highest coefficient in most endpoints and the gradient coefficient of the DDscore (**Figure S5**).

Adding the DDscore to the base model significantly improved predictive information over the base model for most AA and AAA, but for IA endpoints (**Figure 3b**, **Figure S6, Table S16**). The addition of DDscore didn’t enhance the base model’s predictive capability for unruptured AAA. However, DDscore markedly boosts the ability to predict ruptured AAA (C-index [95%CI)]: Proteins + DDscore 0.93 [0.88-0.99] and Proteins + RF+ PRS + Metabolites+ DDscore 0.94 [0.91-1.00]). For all AA events, DDscore could improve the predictive ability of the base models, including AA (Proteins + DDscore 0.79 [0.76-0.83] and Proteins + RF+ PRS + Metabolites+ DDscore 0.82 [0.79-0.85]), unruptured AA (Proteins + DDscore 0.78 [0.75-0.81] and Proteins + RF+ PRS + Metabolites+ DDscore 0.80 [0.77-0.85]) and ruptured AA (Proteins + DDscore 0.92 [0.86-0.99] and Proteins + RF+ PRS + Metabolites+ DDscore 0.93 [0.89-0.96]). For participants with a higher percentile of model risk score at baseline, integrating DDscore exhibited a higher prevalence for the top 10% of both ruptured AAA and unruptured AA than the base model (**Figure S4b**). After integrating DDscore, the Kaplan-Meier survival curves showed more distinctive paths between the tertiles stratified than base model for most AA and AAA, but for IA endpoints (**Figure S7**, **Table S13**). For predictive abilities across different time windows in most AA and AAA endpoints, DDscore could enhance the AUC, yet it couldn’t alter the fact that the peak AUC was still predicting outcomes within 5 years (**Table S14**). For seven of the nine endpoints, DDscore integrated models achieved higher LRs than base models at a 10% false positive rate (**Figure 3b**, **Table S4**). Notably, the LR improvement was noteworthy for AA and AAA, especially for ruptured AAA (ΔLR = 1.18 for Proteins + DDscore compared to Proteins; ΔLR = 0.53 for Proteins + RF+ PRS + Metabolites+ DDscore compared to without DDscore). For all events, the models integrating RF + PRS + Metabolites had an average net reclassification improvement (NRI) of 0.046 (25th–75th percentile = 0.029–0.043), while the mean NRI across DDscore integrated models was only 0.007 (25th–75th percentile = 0.003–0.012; **Table S17-S18**). However, DDscore outperformed the integrated models with greater NRI for ruptured AAA.

### Discriminative performance translates to model calibration and clinical utility

While discrimination is critical, the clinical utility of any risk model depends on calibration and the choice of adequate thresholds for interventions. We assessed the predictive models by examining calibrations and performing decision curve analysis on the testing set. Models for almost all AA and AAA endpoints were well calibrated, where the observed risk and the predicted risk showed consistency, except for IA (**Figure 4a, Figure S8a**, **Table S19**). The net benefit curves showed the trade-offs of benefits and harms for clinical decisions based on different models across a range of decision thresholds (**Figure S8b**). For all endpoints, the Proteins + RF + PRS + Metabolites showed greater net benefit than other base models. For all AA and AAA endpoints, the Proteins outperformed other single-modality models. Within certain decision thresholds, the Proteins even demonstrated a higher net benefit than the Proteins + RF + PRS + Metabolites in AA and AAA rupture. The DDscore integrated models showed great performance in discriminating populations at risk, but such results could not implicate whether DDscore should be used in clinical practice. To determine the enhancement of clinical utility through the integration of DDscore, we evaluated the performance of integrated models to uncover potential net benefits between them (**Figure 4b**). The addition of DDscore to Proteins or Proteins + RF + PRS + Metabolites substantially improved the clinical utility. For all events, the Proteins + RF + PRS + Metabolites + DDscore Model showed greater net benefit compared with other models. Compared to the Proteins + RF + PRS + Metabolites + DDscore Model, the Proteins or Proteins + DDscore models offer greater net population benefit within the 0%-2% decision threshold for ruptured AA and 0%-1% for ruptured AAA.

**Figure 4.**
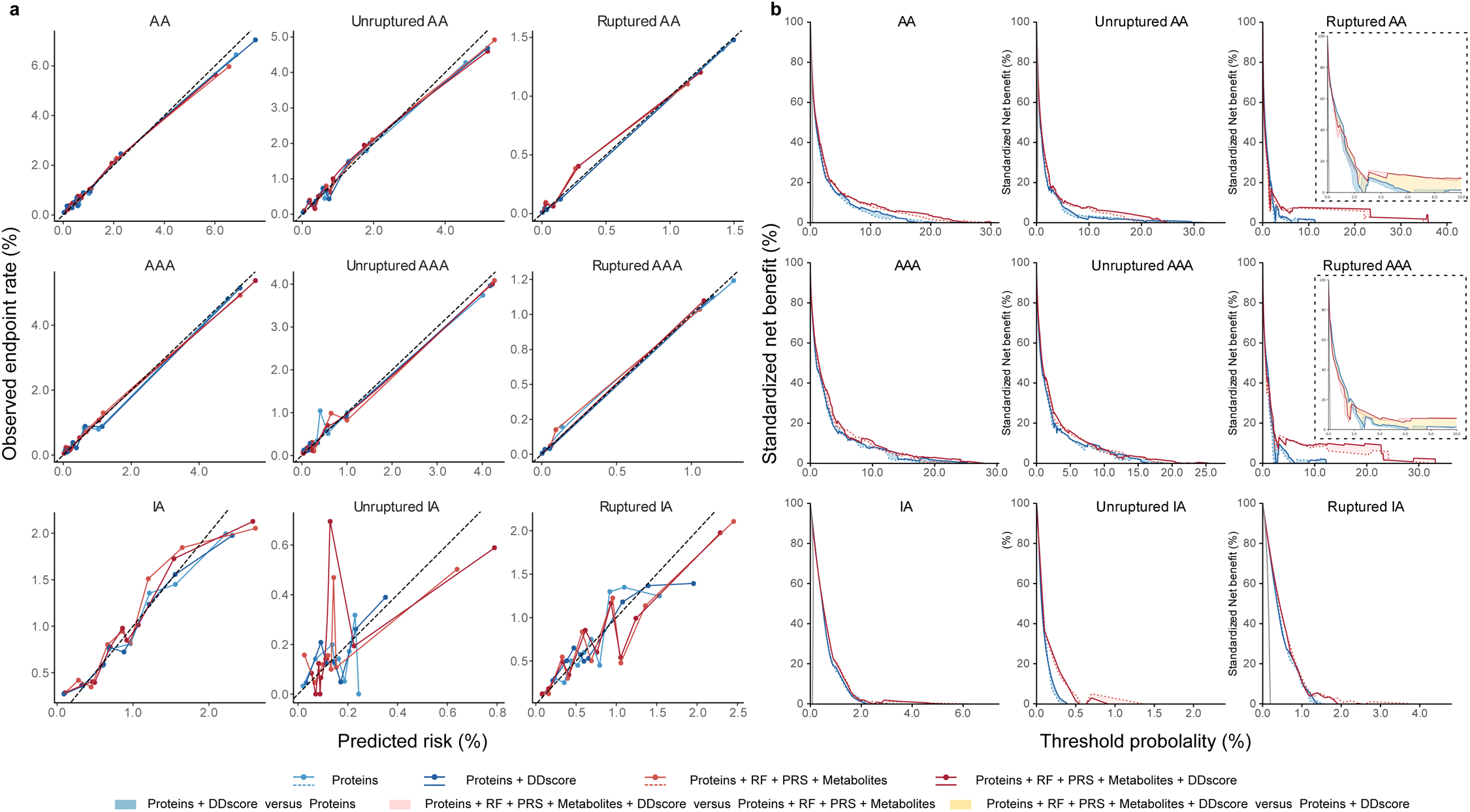
Model calibration and the additive predictive value of proteins and DDscore demonstrate potential clinical utility. **a,** Calibration curves for CPH models trained on Proteins (blue), Proteins + DDscore (dark blue), Proteins + RF + PRS + Metabolites (Four-modality, red), and Proteins + RF + PRS + Metabolites + DDscore (dark red) across nine aneurysm endpoints. **b,** Standardized net benefits of the sets Proteins, Proteins + DDscore, Proteins + RF + PRS + Metabolites (Four-modality), and Proteins + RF + PRS + Metabolites + DDscore for all endpoints. Blue, pink, and yellow shaded areas indicate the added benefit of combining Proteins with Proteins + DDscore, Four-modality with Four-modality + DDscore, and Proteins + DDscore with Four-modality + DDscore, respectively.

### Identification of specific biomarkers in aneurysm events

Compared to the other three single-modality base models, the Proteins Model exhibits better predictive performance and higher cost-effectiveness across most endpoints, particularly in AA and AAA rupture. Moreover, the Proteins Model almost always holds the most significant weight in all integrated models (**Figure S5**, **Table S20**). To understand individual proteins in the context of the nine investigated endpoints, we examined global proteomics attributions by applying Max-Normalization to the non-zero coefficients obtained after fitting the LASSO regression model (**Figure 5a**, **Table S4**). FASLG, MMP12, EGFR, CNTN1, and CEACAM5 demonstrated important predictive value across over half of the endpoints. Among them, MMP12 played crucial roles in the prediction of all events of AA and AAA, while FASLG is the highest-weight predictive factor in IA rupture, showing consistency with the findings reported in other studies^38–40^. Some proteins could predict unruptured aneurysms and were also better at predicting ruptured aneurysms. Therefore, we inferred that these proteins are related to the severity of the occurrence and development of aneurysms, including MMP12 and FASLG for AA, and MMP12, TNR, and CNTN1 for AAA. Elevated plasma levels of NRP1, SIGLEC5, and HS6ST2 were identified to an increased risk of unruptured AAA, while higher concentrations of TIMD4, ZNF830, and ICAM5 in plasma were connected to a greater risk of unruptured IA. Notably, the greatest value of the Proteins Model was in its prediction of aneurysm rupture, significantly surpassing other base models. For this reason, we have confirmed several proteins with high-weight specific predictive abilities for aneurysm rupture: KLKB1, PZP, and RARRES1 for AA rupture; INSL3, OMP, and GALNT3 for AAA rupture; and NEFL, SERPINB5, and VSIG4 for IA rupture (**Figure 5b, Figure S9a**). We also standardized the contribution of metabolites in the Metabolites Model (**Figure S9b, Figure 13)**. High levels of glutamine, tyrosine, and citrate were observed to have risk effects associated with IA rupture. Additionally, total fatty acids, the overall concentration of lipoprotein particles, and phospholipids present in HDL were identified as possible factors contributing to the risk of unruptured AA and AAA. Detailed data regarding all the endpoints examined can be found in **Table S3**.

**Fig. 5.**
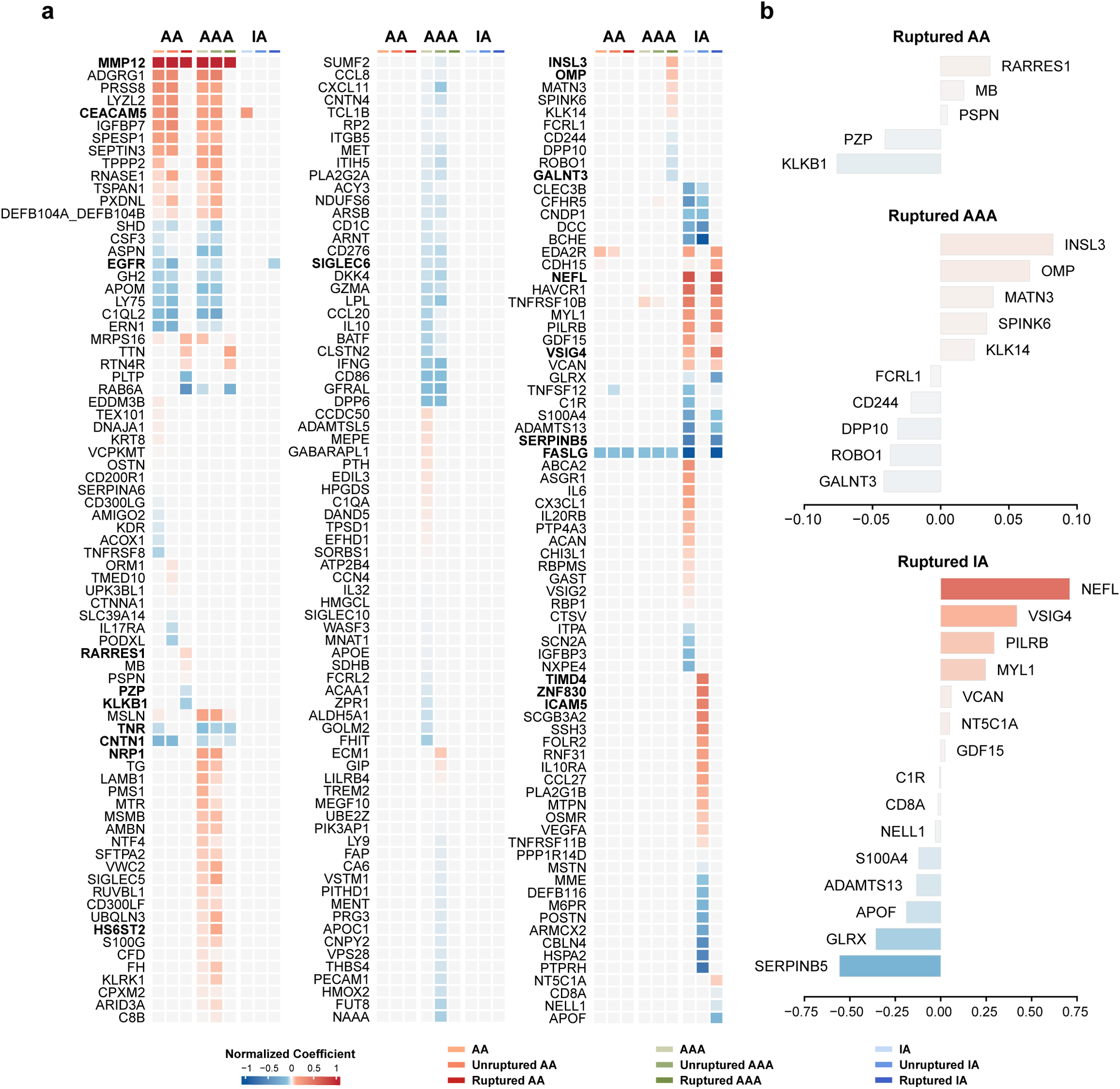
Analysis and attributions of the plasma proteomic profiles associated with aneurysm risk. **a,** Heatmap showing the importance of plasma proteins regarding the Proteins Model for nine endpoints, represented by normalized coefficients, visualized the identification of aneurysm-specific proteomic profiles (Table S4). **b,** Specific protein attributions for ruptured aneurysm events, represented by normalized coefficients. The proteins mentioned in the manuscript have been bolded.

## Discussion

Aneurysms typically enlarge asymptomatically until intervention through either open or endovascular surgery or until rupture, which can result in catastrophic outcomes, including death^3,7,41^. Early prediction is critical for identifying high-risk populations for aneurysm incidence and rupture. Existing studies have developed models using clinical and genomic data for prediction, but their performance is inconsistent^12,13,18^. Predicting the worst outcomes, such as aneurysm rupture, remains particularly challenging without imaging data^21,22^. The recent availability of comprehensive, multi-modality data from over 500,000 individuals in the UK Biobank (UKB) presents an unprecedented opportunity. We developed single- and multi-modality models to predict aneurysm events over a 14.8-year follow-up period using plasma proteomics, metabolomics, genomics, and clinical predictors. Notably, our analysis demonstrated that plasma proteomics could effectively predict most aneurysm outcomes, especially rupture, and outperform other single-modality models. The predictive power of the integrated multi-modality model is only marginally superior to the proteomics model alone. Additionally, leveraging the time-dependent correlations between diabetes and aneurysm events, we developed the Diabetes Duration Score (DDscore) using XGBoost to enhance model performance.

In our single-modality models, we found that PRS was stable but moderately effective; metabolomics slightly outperformed PRS but lacked stability; RF showed good overall performance but struggled with rupture prediction; and proteins demonstrated exceptional predictive power and developmental potential. To our knowledge, this study provides the first comprehensive importance ranking of plasma proteins for predicting future aneurysm events across 5-year, 10-year, over 10-year, and all-time scenarios. Consistent with prior research, proteins such as MMP12^42^ and IGFBP7^43^ have associations with AA and AAA. Moreover, we found that IFNG^44^, APOC1^45^, THBS4^46^, APOM^47^, and NRP1^48^ are correlated with AAA, while FASLG^49^, TNFSF12^50^, ADAMTS13^50^, VCAN^51^, and VSIG4^52^ are linked to IA. These results align with previous findings, highlighting specific aneurysm associations. Additionally, MMP12 and FASLG may serve as indicators of aneurysm severity, predicting both ruptured and unruptured events. Unique to this study, we identified novel associations between specific proteins and ruptured aneurysms: KLKB1, RARRES1, and PZP with ruptured AA; INSL3, OMP, and GALNT3 with ruptured AAA; and NEFL, PILRB, and SERPINB5 with ruptured IA. These proteins may be involved in aneurysm pathogenesis via vascular remodeling^42^, immune response^37^, inflammation^28^, and vascular smooth muscle cell phenotypic switching^48^. However, further research is needed to elucidate the mechanisms of these rupture-specific proteins.

Diabetes is a known risk factor for cardiovascular disease^16^, but its effect on aneurysm incidence and rupture has been paradoxical in epidemiological studies^14,15^. Single-modality data yielded inconsistent or non-significant relationships between HbA1c, glucose, and aneurysm outcomes, complicating the assessment of hyperglycemia’s impact. We observed that antidiabetic medications were protective against AA and AAA incidence^50,54^, but not significantly associated with rupture^55^, consistent with other studies. Prevalent diabetes at enrollment did not protect against aneurysms (*p* > 0.05). In contrast, post-enrollment diabetes incidence was a risk factor for seven endpoints. By defining diabetes duration, we clarified these inconsistent associations, revealing a stable time-dependent correlation: shorter duration of diabetes was associated with fewer aneurysm events, while longer duration increased them. This time-dependent correlation warrants further investigation and may guide clinical aneurysm risk management

This study developed an enhanced predictive model, the DDscore, based on observed time-dependent correlations. For AA and AAA endpoints, the DDscore significantly boosts the predictive accuracy of the base model, though this improvement is less evident for IA endpoints. For the first time, our model surpassed 0.9 in predicting aneurysm rupture risk without relying on CTA or MRI. In the Proteins + RF + PRS + Metabolites + DDscore model, the peak C-index for ruptured AA reached 0.93 [0.89–0.96], while for ruptured AAA, it achieved 0.94 [0.91–1.00]. Even with just Proteins + DDscore, the C-index was 0.92 [0.86–0.99] for ruptured AA and 0.94 [0.90–0.99] for ruptured AAA. Predictive performance generally declined as the follow-up period lengthened. We suggest that individuals flagged by this model as high-risk for rupture should receive timely intervention and preventative care. Randomized controlled trials are essential to establish evidence-based guidelines for aneurysm surgery.

Our study has several key strengths, being the first to combine proteomics and metabolomics data with comprehensive aneurysm endpoint analysis, especially for rupture. We identified numerous biomarkers and highlighted the value of incorporating diabetes duration to enhance predictive models. Supported by one of the world’s largest and longest prospective cohort studies, our findings allow for precise clinical assessment. However, certain limitations warrant consideration. First, our sample included only individuals with aneurysms identified via ICD codes, excluding undiagnosed cases and lacking aneurysm size data, constraining model optimization. Second, despite the large UKB cohort, the low incidence of certain outcomes caused by undetected asymptomatic aneurysms affects model stability. Third, as most UKB participants are of white European descent, we did not account for race/ethnicity, which could influence risk prediction. Lastly, large-scale, multi-modality data with long follow-ups remain scarce. While internal cross-validation produced well-calibrated models, external validation in diverse primary care settings is essential prior to clinical implementation.

## Conclusion

Plasma proteomics showed exceptional predictive advantages over other single-modality data, outperforming standard clinical predictors for aneurysm events, particularly rupture. Furthermore, incorporating the time-dependent correlation between diabetes and aneurysm risk further improved model accuracy, reclassification, and clinical relevance. In summary, we developed an imaging-free predictive model for future aneurysm events, emphasizing the utility of proteomics and diabetes duration in clinical practice to identify high-risk individuals and support early intervention.

## Acknowledgements

We extend our gratitude to all the researchers and participants who contributed to the UK Biobank.

## Data availability

This research used data from UK Biobank, a major biomedical database, via application number 89885. UK Biobank data are publicly available to bona fide researchers upon formal application at https://www.ukbiobank.ac.uk/. Detailed information about the data used in this study, including participant characteristics and outcomes, can be found in the Supplementary Information.

## Code availability

All software used in this study is publicly available. The code developed and used throughout this study has been made open source and can be accessed at https://github.com/RenBichen/AneurysmPrediction.

## Author contributions

B.R. was responsible for the conception, design, and overall supervision of the project. B.R. and L.Y. drafted the manuscript and organized all tables and figures, and J.W. verified the underlying data reported in the manuscript. T.X. conducted basic data cleaning, implemented the development of the machine Learning models, and performed the statistical analyses. Q.Z., X.Y., and B.R. supplemented and revised the model construction and analysis and contributed to interpreting the interpretable machine learning results and supported model validation. X.C, Q.Z., and X.Y. provided analytical support and contributed to the discussion of the results. Y.F., T.P., G.F., and L.C. provided recommendations on the clinical applicability. Z.D., M.W., J.W., and W.F. revised critically for important intellectual content and approved the final version of the manuscript. All authors accepted responsibility for submitting the manuscript for publication.

